# Ultra-rapid on-site detection of SARS-CoV-2 infection using simple ATR-FTIR spectroscopy and analysis algorithm: high sensitivity and specificity

**DOI:** 10.1101/2020.11.02.20223560

**Authors:** Valério G. Barauna, Maneesh N. Singh, Leonardo Leal Barbosa, Wena Dantas Marcarini, Paula Frizera Vassallo, Jose Geraldo Mill, Rodrigo Ribeiro-Rodrigues, Patrick H. Warnke, Francis L Martin

## Abstract

There is an urgent need for ultra-rapid testing regimens to detect the SARS-CoV-2 [Severe Acute Respiratory Syndrome Coronavirus 2] virus infections in real-time within seconds to stop its spread. Current testing approaches for this RNA virus focus primarily on diagnosis by RT-qPCR, which is time-consuming, costly, often inaccurate and impractical for general population rollout due to the need for laboratory processing. The latency until the test result arrives with the patient has led to further virus spread. Furthermore, latest antigen rapid tests still require 15 to 30 min processing time and are challenging to handle. Despite increased PCR-test and antigen-test efforts the pandemic has entered the worldwide second stage. Herein, we applied a superfast reagent-free and non-destructive approach of attenuated total reflection Fourier-transform infrared (ATR-FTIR) spectroscopy with subsequent chemometric analysis to the interrogation of virus-infected samples. Contrived samples with inactivated gamma-irradiated Covid-19 virus particles at levels down to 1582 copies/ml generated infrared (IR) spectra with good signal-to-noise ratio. Predominant virus spectral peaks are associated with nucleic acid bands, including RNA. At low copy numbers, the presence of virus particle was found to be capable of modifying the IR spectral signature of saliva, again with discriminating wavenumbers primarily associated with RNA. Discrimination was also achievable following ATR-FTIR spectral analysis of swabs immersed in saliva variously spiked with virus. Following on, we nested our test system in a clinical setting wherein participants were recruited to provide demographic details, symptoms, parallel RT-qPCR testing and the acquisition of pharyngeal swabs for ATR-FTIR spectral analysis. Initial categorisation of swab samples into negative *versus* positive Covid-19 infection was based on symptoms and PCR results. Following training and validation of a genetic algorithm-linear discriminant analysis (GA-LDA) algorithm, a blind sensitivity of 95% and specificity of 89% was achieved. This prompt approach generates results within two minutes and is applicable in areas with increased people traffic that require sudden test results such as airports, events or gate controls.

## Introduction

In early 2020, a new strain of coronavirus called SARS-CoV-2 (Severe Acute Respiratory Syndrome Coronavirus 2), more commonly known as causing the Covid-19 disease, gave rise to a global pandemic (1). Starting from an epidemic outbreak in Wuhan (China), the virus quickly spread westwards towards Europe and the USA (2) with serious health and socio-economic consequences worldwide (3). SARS-CoV-2 exhibits a high propensity for infectious spread throughout populations (2). Every Covid-19 positive case, if not contained, can readily spread to two or more people giving a virulent R number (4). Some countries, such as South Korea, initially successfully fought the Covid-19 outbreak. This is based on the key aspects (5) of: (a) prevention, *via* good cleaning practices and isolation of potential cases; (b) testing, to identify those infected and to precisely isolate risk cases; and, (c) anti-viral treatment and, in the future, a vaccine. Testing is fundamental to identify infected people and regions of risk (6). This can enable intelligent isolation of areas without affecting an entire country’s economy and allow allocation of resources to more strategically fight the disease, with more ventilators, medication and medical staff assigned to regions with more diagnosed cases.

The main challenges for testing are the cost and in particular time. Gold-standard diagnosis by RT-qPCR is costly with a shortage of testing facilities even in developed countries and can take >2 days to get the result, because specimen have to be transported for processing to often distant laboratories (7). This is not suitable for mass testing (8). Despite globally increased PCR-test efforts the pandemic was not brought to a halt. In contrast, there is a recurrence and second wave of the disease, because many infectious patients spread the disease while waiting on their PCR-test results. There are some companies are developing quicker and lower-cost tests based on novel sensors (9). Alternative antigen- or antibody-detection approaches remain affected by low specificity (that is, healthy patients could be wrongly classified as Covid-19 positive) thus creating statistical bias that could directly affect public health policies (10). Thus, there is a need to develop COVID-19 test approaches that can deliver results in real time and on-site.

Vibrational spectroscopy, including attenuated total reflection Fourier-transform infrared (ATR-FTIR) spectroscopy, has been widely used to discriminate and classify normal and pathological populations using different cell types, tissues or biofluids (11-13). Readily accessible biofluids, such as blood plasma/serum, saliva or urine, are considered ideal for clinical implementation due to routine methods of collection, as well as minimal sample preparation (14). Interrogation of samples with infrared (IR) spectroscopic techniques allows for the generation of a “spectral fingerprint” which subsequently facilitates the discrimination of the different populations and identification of potential biomarkers (15). In the past few years, biofluid-based ATR-FTIR spectroscopy have been used for diagnosing, screening or monitoring the progression/regression in a variety of diseases (16). Spectroscopic techniques are rapid, cost-effective and non-destructive which make them a perfect candidate for translation to clinic.

As a readily accessible non-invasive biofluid, saliva is an ideal candidate to facilitate disease detection; indeed, oral health has long been known to be an indicator of whole organism health (17). Herein, ATR-FTIR spectroscopy was used to interrogate saliva samples on pharyngeal swabs taken from individuals with or without suspected infection with Covid-19. Unlike many tests developed using laboratory-based contrived specimens, we trialled the approach in clinical settings on real-world samples. Our goal was to differentiate individuals with active infection based on a series of spectral biomarkers. We also took into consideration symptoms and other demographic features of our participants as confounding factors. We propose a new, ultra-rapid on-site method to detect Covid-19 based on pharyngeal swabs using IR light, with potential for ready implementation in general population settings.

## Results and Discussion

### Spiking of saliva with inactivated Covid-19 virus

Figure 1a shows a typical spectrum of inactivated gamma-irradiated Covid-19 virus particles (e SARS.CoV2/SP02.2020.HIAE.Br GenBank accession number MT 126808.1) (18); at 1582 copies/ml, an ATR-FTIR spectrum with good signal-to-noise ratio (SNR) is obtained. This was in order to assess the limit of detection (LoD) for biospectroscopy to ascertain the minimum concentration at which the virus could be detected by IR spectroscopy. Below this level, the SNR becomes poor and noisy. This clearly points to the ability of ATR-FTIR spectroscopy to extract a unique viral fingerprint consistent of spectral features associated with a pure virus spectrum. It is interesting to note that the predominant spectral peaks are associated with nucleic acid bands, including RNA. Following nucleic acid (RNA/DNA) extraction of saliva samples obtained from participants either positive (*n*=5) or negative (*n*=5) for Covid-19, a clear segregation of spectral data points is obtained using exploratory principal component analysis (PCA) (Figure 1b).

**Figure 1.**
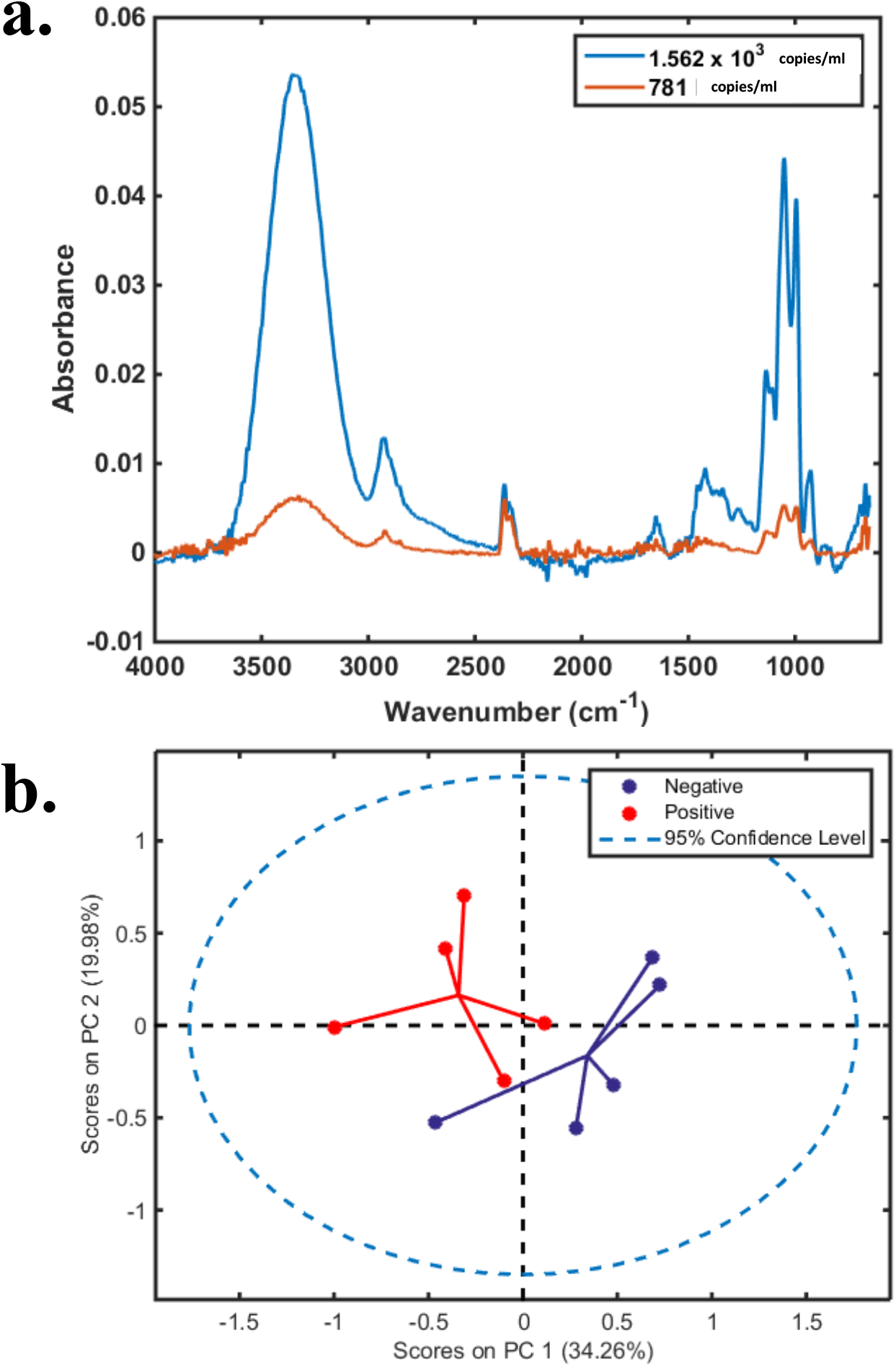
a. Spectra of pure virus in saliva (whole virus inactivated by Gamma-radiation) (e SARS.CoV2/SP02.2020.HIAE.Br GenBank accession number MT 126808.1). b. Graphical demonstration of separation of positive and negative sample in RNA extracted samples prepared for PCR analysed by biospectroscopy.

Control saliva from a human participant (Male, 42 y and RT-qPCR negative) was spiked with various numbers of inactivated gamma-irradiated Covid-19 virus particles (Figure 2). At low copy numbers, the virus particle is clearly capable of modifying the infrared spectral signature of saliva (Figure 2a and 2b). Examination of control saliva in comparison with saliva spiked inactivated virus particle at various copy number levels highlighted an ability to detect virus particle-induced spectral alterations at levels that would be considered extremely low in the pharyngeal cavity of infected humans (symptomatic or asymptomatic). Even more compellingly, when this is examined using basic multivariate analysis (*i*.*e*., PCA), the IR spectral signature of pure inactivated virus segregates away from control saliva in a scores plot (Figure 2c). When saliva is spiked with exceptionally low levels of virus (781 copies/ml; a1 cluster below), the spectral points co-cluster with control saliva spectral points suggesting no differences. However, at a level of 12,500 copies/ml (a2 cluster below), there is segregation away from the control. It is critical to note that the loadings plot specifically identifies RNA as being proportional to virus levels (Figure 2b). The loadings on PC1 show the bands responsible for increase of virus concentration (nucleic acid bands), and the loadings on PC2 shows the bands responsible for discrimination between saliva and virus (Amide I and Amide II bands present in saliva but not virus). Other, primarily protein-associated bands discriminate the saliva from the virus – we believe this to be the first report of its kind using biospectroscopy.

**Figure 2.**
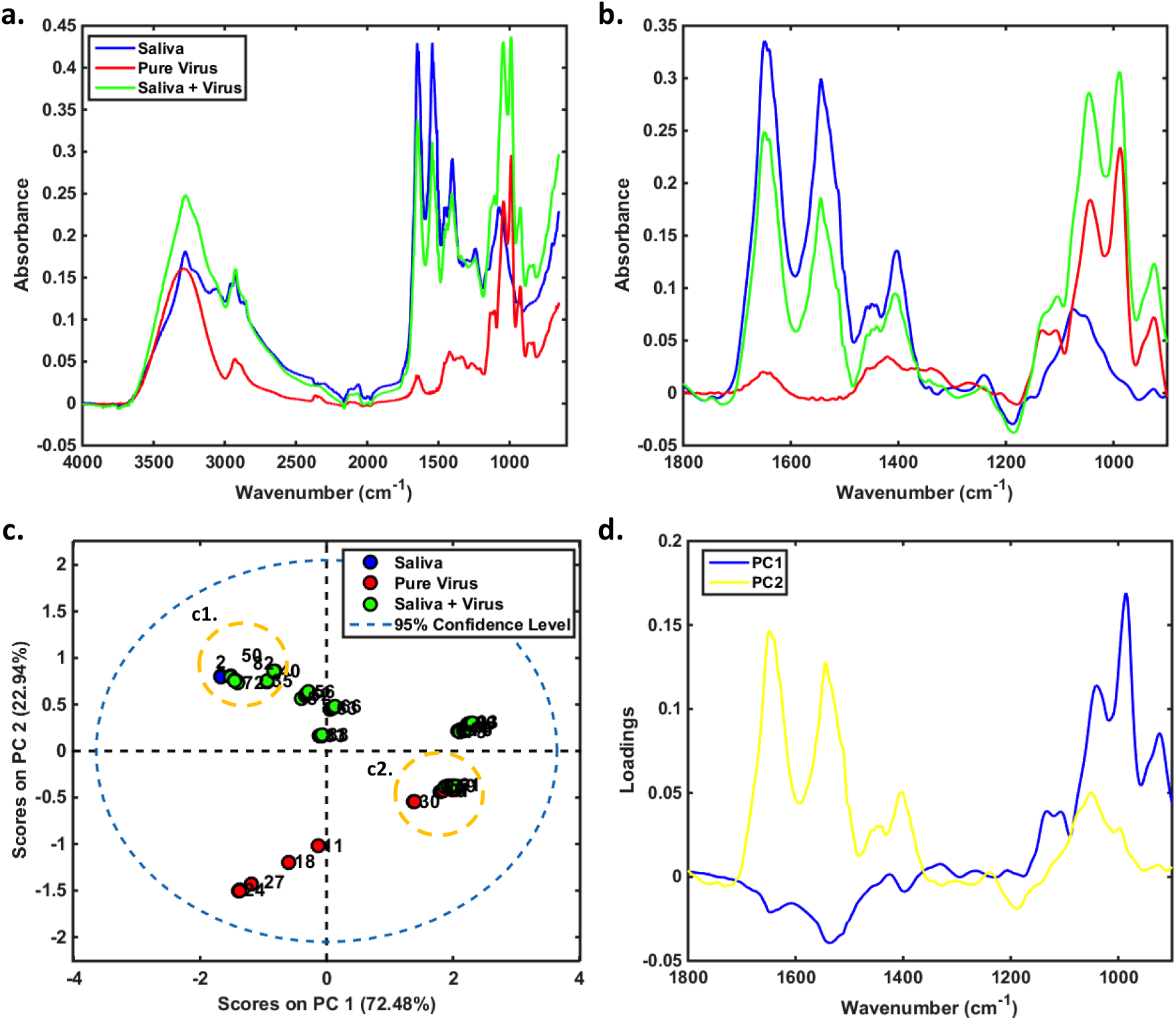
(a) Average raw spectra and (b) pre-processed spectra for Saliva (*n* = 2), Pure Covid-19 Virus in different concentrations (*n* = 28, 1×10^5^-98 copies/mL), and Saliva + Virus in different concentrations (*n* = 63, 1×10^5^-24 copies/mL). (c) PCA scores and (d) PCA loadings on PC1 *vs*. PC2 for the pre-processed data. Inset c1.: Mix between saliva and saliva + virus for low concentration (≤ 781 copies/mL); c2.: Mix between pure virus and saliva + virus for high concentration (≥ 1.25×10^4^ copies/mL). Pre-processing: Savitzky-Golay (SG) smoothing (7 point window, 2^nd^ order polynomial fitting) and baseline correction. The loadings on PC1 show the bands responsible for increase of virus concentration (nucleic acid bands), and the loadings on PC2 shows the bands responsible for discrimination between saliva and virus (Amide I and Amide II bands present in saliva but not virus).

Furthermore, in the complex milieu of a saliva sample, which will undoubtedly contain a range of complex constituents including aqueous, exfoliated cellular material, post-infection immunoglobulins such as IgA and other individual or contaminating factors, a multivariate chemometric approach can still extract the viral-associated discriminating features. Following this, Figure 3 shows the analysis of swabs spiked with either saliva with or without spiking with gamma-irradiated Covid-19 virus particles. Figures 3a and 3b show spectra with good SNR. In consequent PCA scores plots, the spectral data points for virus-spiked saliva swabs segregate away from swab or control saliva swab categories (Figure 3c). This is achieved at low copy numbers. The loadings on PC1 show the bands responsible for separation between swab + saliva and swab + saliva + virus (Amide I and Amide II band of proteins) and the loadings on PC2 show the bands responsible for variation of virus concentration (Amide I, Amide II and nucleic acids bands) (Figure 3d). Differently from saliva, the swab sample contains bands on the nucleic acids region plus Amide I and Amide II that may come from the saliva itself.

**Figure 3.**
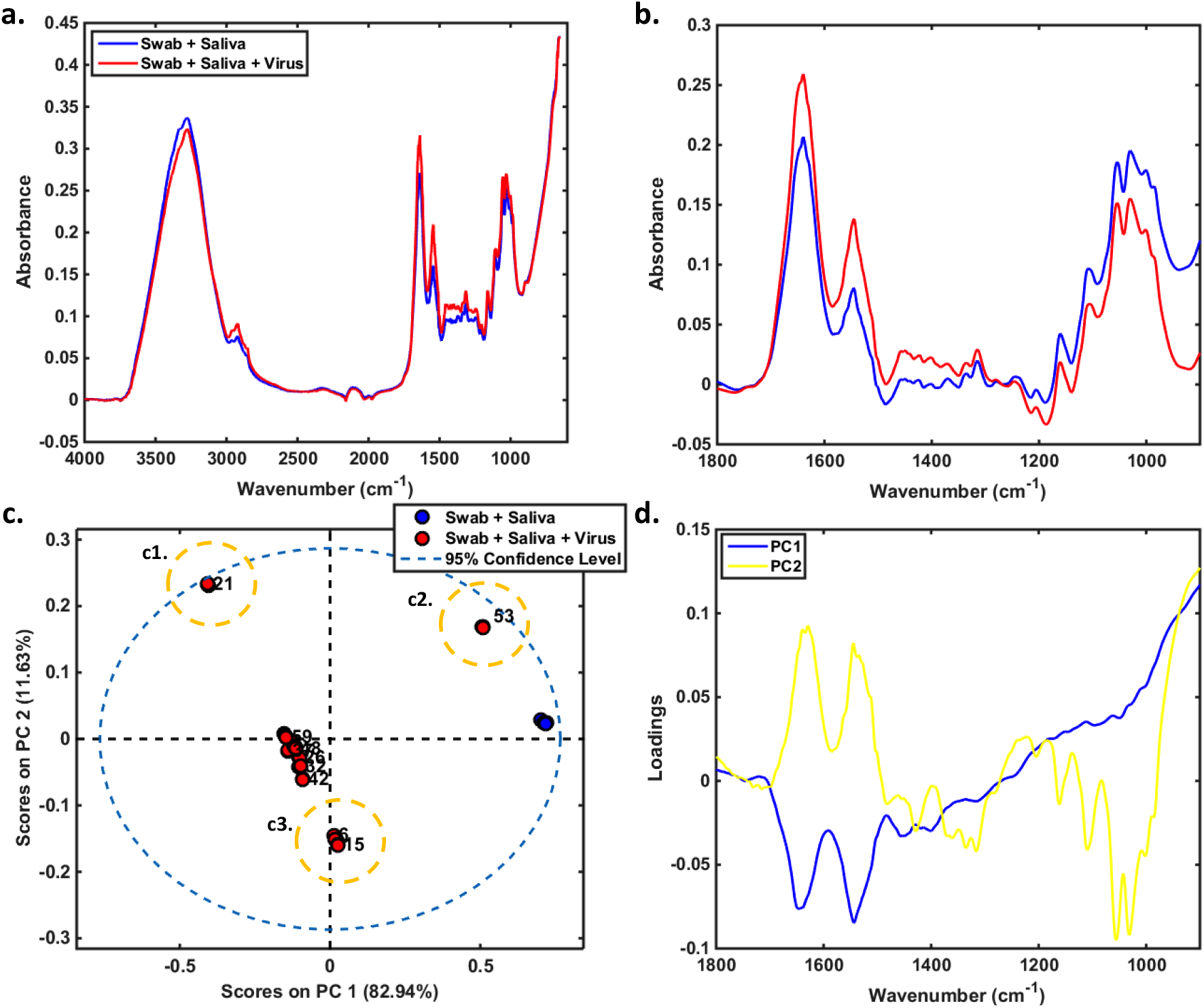
(a) Average raw spectra and (b) pre-processed spectra for Swab + Saliva (*n* = 5), and Swab + Saliva + Virus (*n* = 54, 1×10^5^-98 copies/mL). (c) PCA scores and (d) PCA loadings on PC1 *vs*. PC2 for the pre-processed data. Inset c1.: virus concentration around 6.25×10^3^ copies/mL. c2.: virus concentration around 1.56×10^3^ copies/mL; c3.: virus concentration ≤ 781 copies/mL. Pre-processing: Savitzky-Golay (SG) smoothing (7 point window, 2^nd^ order polynomial fitting) and baseline correction. The loadings on PC1 show the bands responsible for separation between swab + saliva and swab + saliva + virus (Amide I and Amide II band of proteins) and the loadings on PC2 show the bands responsible for variation of virus concentration (Amide I, Amide II and nucleic acids bands). Differently from Saliva, the Swab sample contains bands on the nucleic acids region plus Amide I and Amide II that may come from the saliva itself.

### GA-LDA segregation of categories: Covid-19 infected vs. uninfected

Categorisation into the negative (designated not infected by Covid-19) and positive (designated as infected) categories was based on a series of RT-qPCR tests including the Berlin protocol (18) primarily carried out at Central Laboratory of Espírito Santo State alongside symptoms/outcome. Follow-up showed that Covid-19 participants required hospitalisation. A Ct <37 in RT-qPCR designated a PCR-positive result (Figure 4).

**Figure 4.**
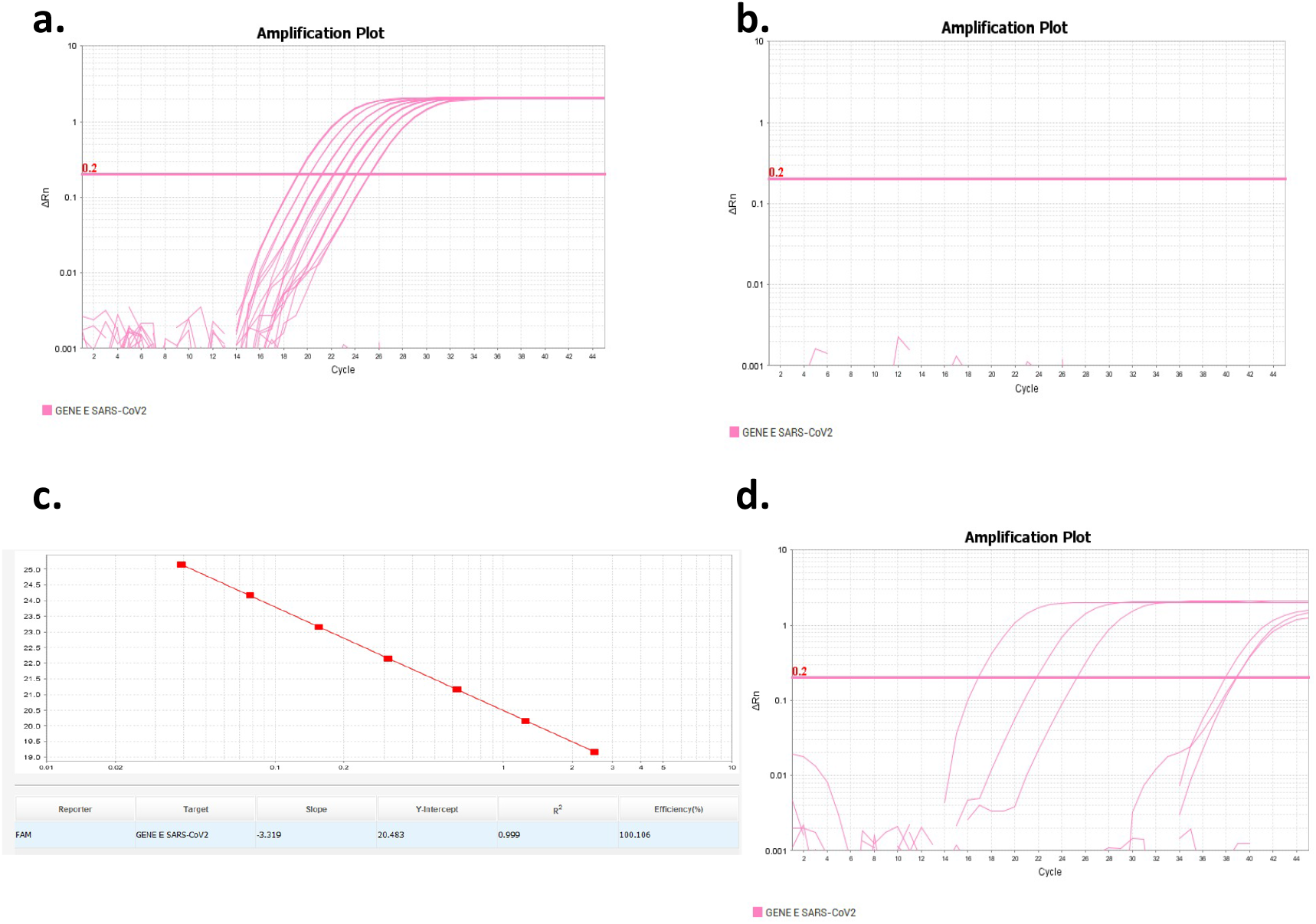
qRT-PCR of samples. A tiered RT-qPCR system including the Berlin protocol was employed for the analyses of parallel naso-pharyngeal samples taken from study participants. **a**. The samples were considered positive if the E gene amplified with Ct <37. **b**. A standard curve is below alongside a negative control. **c**. The efficiency curve to determine the threshold is shown. **d**. Three positive samples are shown juxtaposed with three negative samples (no amplification) and three with a Ct >37 (negative/inconclusive).

The chemometric technique of genetic algorithm-linear discriminant analysis (GA-LDA) was applied towards classification (19, 20) of negative *versus* positive for Covid-19 infection (Figure 5). The classification ratios achieved after GA-LDA were a sensitivity of 95% and specificity of 89% (Table 1). Figure 5a and 5b shows the full raw spectra across the entire mid-IR spectral range and raw spectra in the fingerprint region for all negative and Covid-19 positive swab samples (*n* = 111 negatives and 70 positives). Spectra were pre-processed [Savitzky-Golay smoothing (9 point window, 2^nd^ order polynomial fitting), automatic weighted least squares baseline correction and vector normalisation] in the fingerprint region (Figure 5c). Training and validation of GA-LDA was undertaken using 50 negatives and 50 positives; the GA-LDA scores plot for the validation set (*n* = 61 negatives and 20 positives) is shown in Figure 5d.

**Table 1.** Confusion matrix and figures of merit for the validation set using GA-LDA algorithm pre-processing.

| ***Baseline correction and vector normalisation\**** |  |  |
| --- | --- | --- |
|  | Predicted Negative | Predicted Positive |
| Negative | 54 | 7 |
| Positive | 1 | 19 |
| **Parameters** |  |  |
| Accuracy | 90% |  |
| Sensitivity | 95% |  |
| Specificity | 89% |  |
| F-Score | 92% |  |

\*Pre-processing: Savitzky-Golay smoothing (9 point window, 2<sup>nd</sup> order polynomial fitting), automatic weighted least squares baseline correction and vector normalisation.

**Figure 5.**
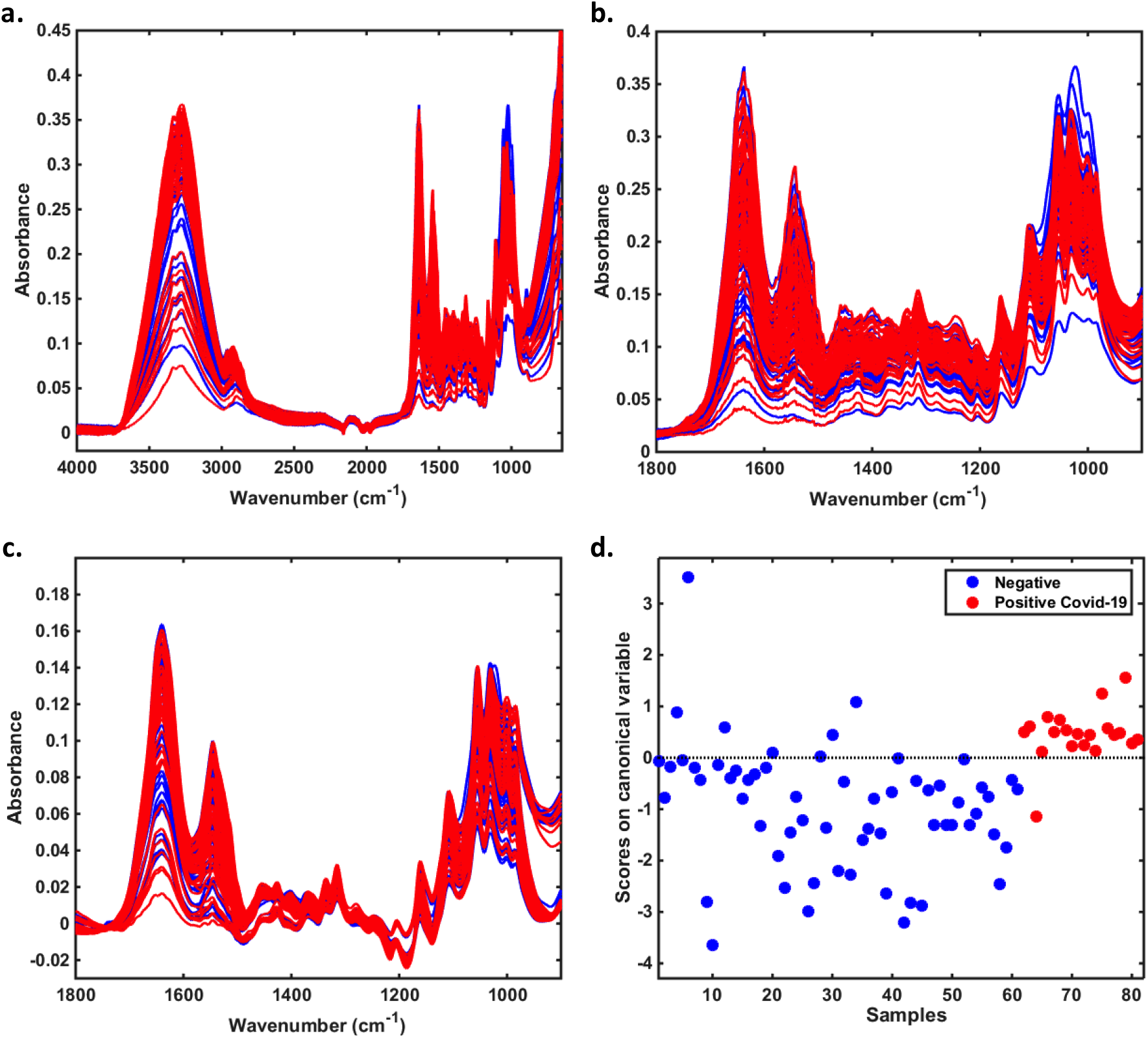
(a) Full raw spectra and (b) raw spectra in the fingerprint region for all negative and Covid-19 positive swab samples (*n* = 111 negatives and 70 positives). (c) Pre-processed spectra [Savitzky-Golay smoothing (9 point window, 2^nd^ order polynomial fitting), automatic weighted least squares baseline correction and vector normalisation] in the fingerprint region and (d) GA-LDA scores plot for the validation set (*n* = 61 negatives and 20 positives).

Consequently, five GA-LDA selected variables were identified, each which significantly (*P* <0.01) discriminates negative and Covid-19 positive swab samples (Figure 6, Table 2). Using saliva swab-based vibrational spectroscopy we achieved results with significant clinical relevance. ATR-FTIR spectroscopy has been proven to be capable of distinguishing between patient and healthy groups negative and Covid-19 positive swab samples. The plausible mechanistic basis for this is that the prominent distinguishing features extracted are primarily associated with nucleic acids, RNA in particular.

**Table 2.** Selected variables by GA-LDA with their respective tentative assignments.

| **Variable (cm<sup>-1</sup>)** | **Tentative assignment** |
| --- | --- |
| 1429 | $\delta(\text{CH}_2)$ polysaccharides |
| 1220 | Asymmetric $\text{PO}_2^-$ stretching in RNA and DNA |
| 1084 | Symmetric $\text{PO}_2^-$ stretching in nucleic acids |
| 1069 | C-O stretching in ribose |
| 1041 | Symmetric $\text{PO}_2^-$ stretching in nucleic acids |

**Figure 6.**
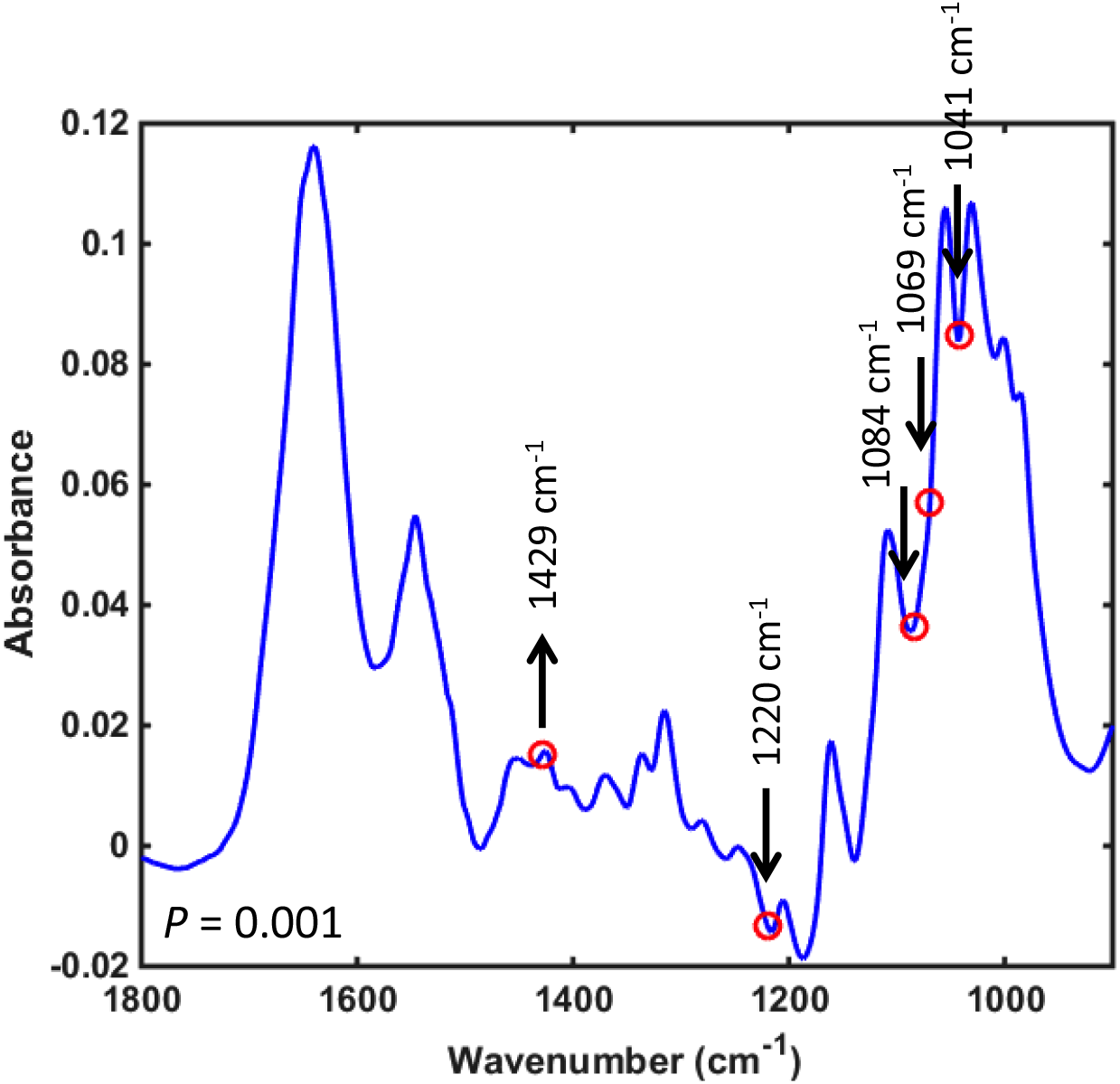
GA-LDA selected variables. Arrow ↑: higher absorbance in the Covid-19 positive class. Arrow ↓: higher absorbance in the Covid-19 negative class. *P*-value calculated using a MANOVA test with all 5 GA-LDA selected variables between all negative and positive samples.

## Methods

### Ethical approval

This study was carried out in agreement with the Helsinki declaration and authorized by the Hospitals Directive, due to the emergency situation. Ethical approval for the investigation was granted by the Ethics Committee Federal University of Espírito Santo (#0993920.1.0000.5071 and #31411420.9.0000.8207). Full ethical approval was given to undertake the studies described herein. All procedures and possible risks were explained to participants before they provided written consent.

### Participant recruitment and swab collection

Pharyngeal cotton swabs (FirstLab, Brazil) were from individuals >18 y, who came to one of the six hospitals participating in the study and met the criteria for suspected cases according to the State Health Secretary and World Health Organization (WHO) guidelines between June and September 2020. For all participants, demographic data (age, gender, pre-existing medical conditions, symptoms, date of symptoms’ onset) were collected. Exclusion criteria were those with inconclusive RT-qPCR results after two rounds of RT-qPCR.

For the gold standard protocol *via* diagnosis by RT-PCR, a nasopharyngeal swab was collected from participants by inserting a rayon swab with a plastic shaft into the nostril parallel to the palate. The swab was inserted to a location equidistant from the nostril and the outer opening of the ear and was gently scraped for a few seconds to absorb secretions. The swab was then placed immediately into a sterile tube containing viral transport medium. RT-PCR was performed in the central laboratory from the Health Secretary of Espírito Santo (LACEN-SESA) to allow definitive diagnosis of COVID-19 infection.

For ATR-FTIR spectroscopy, a pharyngeal swab was collected from participants by inserting a cotton swab into the mouth and scrapping the tonsils, the tongue and the inner part of the cheek. The swab was then placed immediately into a sterile tube and stored on ice until analysis.

### Parallel RT-PCR testing

Samples were taken simultaneously as nasopharyngeal swabs for PCR testing. In the clinical setting, all PCRs were locally or nationally approved tests. All samples were analysed at the same state-approved laboratory.

Nucleic acid extraction and real-time RT-qPCR for virus detection was performed to allow identification of SARS-CoV-2. The extraction of total nucleic acid (DNA and RNA) from collected samples was performed using the BioGene Extraction kit (Bioclin, K204-4, Brazil), following the manufacturer’s instructions. Specimens were handled under the Laboratory biosafety guidance required for the novel coronavirus (2019-nCoV) designated by WHO at the Central Laboratory of the Espírito Santo state (LACEN-ES). A combination of four tests was employed to detect viral RNA. The first was using the IDT (Integrated DNA Technologies; Coralville, Iowa, USA) kit, which is developed in association with the CDC and employs primers and probes for the N1, N2, and RP genes. The second was Maccura (designed by Maccura Biotechnology Co., Hi-tech Zone, Chengdu, China), which is a single-well triple target assay and identifies three genes from SARS-CoV-2 (E, N, and ORF1ab) and provides a separate positive internal control (IC). The third was the Molecular SARS-CoV-2 (E/RP genes) kit (Instituto de Tecnologia em Imunobiológicos, Bio-Manguinhos, FioCruz, RJ, Brazil), which uses primers and probes from the Berlin Protocol (18). The detection of viral RNA was carried out on an ABI 7500 real-time PCR machine (Applied Biosystems, Weiterstadt, Germany), using the published protocol and sequence of primers and probe for E gene and RNAse P. Lastly, the IBMP (Instituto de Biologia Molecular do Paraná – FioCRUZ) kit was employed, which is a single-well test, and detects N and ORF1ab genes, and uses the RP gene as an internal control.

All assays were performed using manufacturers’ recommendations. Firstly, all samples were tested in a single-well assay (IBMP or Maccura) for the qPCR run, and interpretation of all results were added to a spreadsheet, together with the values of Cts obtained. Samples with inconclusive results, either by non-amplification in the internal control or by non-amplification of another gene, were tested with the other two qPCR kits (IDT or Bio-manguinhos gene E). If the PCR result remained inconclusive, the result and Ct values were added to the spreadsheet as negative.

### ATR-FTIR spectral analyses of swabs

FTIR spectra data (wavenumber range 4000–650 cm^-1^) for each swab was obtained by directly placing the saliva swab on a portable Agilent Cary 630 FTIR Spectrometer equipped with an ATR ZnSe crystal (Agilent, Santa Clara, California, United States) and Microlab PC software run from a dedicated computer laptop. Each whole spectrum contains 1798 points (1.86 cm^-1^ spectral resolution). For every ATR-FTIR spectroscopic measurement, three spectra were obtained from each saliva swab. Each swab analysis was performed with 32 co-additions, interspersed with 32 background scans. After each analysis, the swab was removed from the crystal and the crystal was cleaned with miliQ water and 70% alcohol, thus avoiding inter-sample contamination.

### Spiking experiments to determine limit of detection (LoD)

For spiking experiments, gamma-irradiated inactivated Covid-19 virus particles (from a stock solution of 1 × 10^5^ copies/ml deionised water) were mixed in various copy number concentrations in saliva taken from a 42-y-old male classed as negative for infection. The following protocols were undertaken:

1. Four µl of gamma-radiation inactivated Covid-19 virus solution was applied to the ATR diamond and let dry for 4-5 min. Then serial dilutions of the virus in deionized water were analysed in a similar fashion.
2. A series of serial dilutions in saliva from a negative study participant were generated and applied to the ATR diamond and let dry for 4-5 min.
3. Fifteen µl of saliva spiked with gamma-radiation inactivated virus (step 2) were added to a cotton swab. The saliva cotton swab was then applied straight to the ATR diamond and immediately analysed.

### Data pre-processing and analysis

Pre-processing and data analysis were carried out using MATLAB 2014b (The Math Works, MA, USA). The spectra were pre-processed by truncating the fingerprint region (1,800-900 cm^-1^), followed by Savitzky-Golay smoothing (9 point window, 2^nd^ order polynomial fitting), automatic weighted least squares baseline correction and vector normalisation. Towards exploratory data analyses, following pre-processing of raw spectra, spectral data were mean-centred and evaluated by means of principal component analysis (PCA) (21). PCA is an unsupervised technique that reduces the spectral data space to principal components (PCs) responsible for the majority of variance in the original dataset. Each PC is orthogonal to each other, where the first PC accounts to the maximum explained variance followed by the second PC and so on. The PCs are composed of scores and loadings, where the first represents the variance on sample direction, thus being used to assess similarities/dissimilarities among the samples; and the latter represents the contribution of each variable for the model decomposition, thus being used to find important spectral markers. This technique looks for inherent similarities/differences and provides a scores matrix representing the overall “identity” of each sample; a loadings matrix representing the spectral profile in each PC; and a residual matrix containing the unexplained data. Scores information can be used for exploratory analysis providing possible classification between data classes.

PCA was the method of choice for analysing saliva samples spiked with inactivated virus particle. It is simple, fast, and combines exploratory analysis, data reduction, and feature extraction into one single method. PCA scores were used to explore overall dataset variance and any clustering related to limit of detection, while the loadings on the first two PCs were used to derive specific biomarkers indicative of infection category.

Genetic algorithm (GA) is a variable selection technique used to reduce the spectral data space into a few variables and works by simulating the data throughout an evolutionary process (22, 23). The original space is maintained for both algorithms, and no transformation is made as in PCA. Therefore, the selected variables have the same meaning of the original ones (*i*.*e*., wavenumbers), and they are responsible for the region where there are more differences between the classes being analysed or in other words, between the chemical changes.

For all classification models, samples were divided into training (50 designated negative and 50 designated positive for Covid-19 infection based on symptoms and RT-PCR) and validation (*n* = 61 designated negative and 20 designated positive for Covid-19 infection based on symptoms and RT-PCR) sets by applying Kennard-Stone (KS) uniform sampling selection algorithm (19). The training samples were used in the modelling procedure, whereas the prediction set was only used in the final classification evaluation using the LDA discriminant approach. The optimal number of variables for GA was determined with an average risk G of LDA misclassification. Such cost function is calculated in a subset of the training set as:

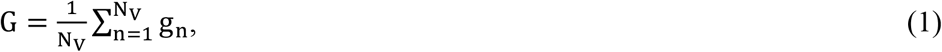

where g_n_ is defined as

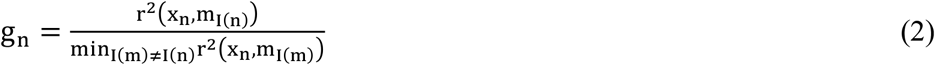

where the numerator is the squared Mahalanobis distance between the object x_n_ and the sample mean m_I(n)_ of its true class; and the denominator is the squared Mahalanobis distance between the object x_n_ and the mean of the closest wrong class (20).

The GA calculations were performed during 100 generations with 200 chromosomes each. One-point crossover and mutation probabilities were set to 60% and 10% respectively. GA is a non-deterministic algorithm, which can give different results by running the same equation/model. Therefore, the algorithm was repeated three times, starting from random initial populations, with the best solution resulting from the three realizations of GA employed.

Sensitivity (probability that a test result will be positive when disease is present) and specificity (probability that a test result will be negative when disease is not present) were given by the following equations:

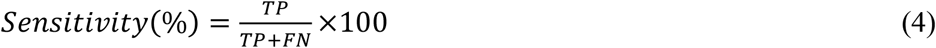

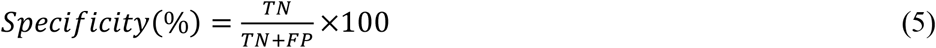

where TP is defined as true positive; FN as false negative; TN as true negative; and FP as false positive.

## Data Availability

All reasonable requests for data will be honoured. The data contained herein in this preliminary is a portion of a much larger study. In due course, when this larger study is published we expect to make the entire dataset available of a publicly accessible data repository.

## Contributions

V.G.B, P.F.V. and J.G.M. were involved in experimental design, writing, review and editing, supervision, project administration and funding acquisition; V.G.B, L.B.L. and W.M. were involved in data acquisition, analysis, and interpretation; M.N.S. and P.H.W. provided clinical expertise, data analytical input and context throughout the course of the study; R.R.-R. undertook and coordinated the RT-qPCR analyses and validation; F.L.M. co-designed the study, was involved in funding acquisition, was principal investigator and wrote the manuscript. All authors approved the finalised manuscript.

## Acknowledgements

We would like to thank LabPetro (UFES, Brazil) for technical support with the ATR-FTIR spectroscopy. We also would like to thank all the hospitals (Vila Velha Hospital, Hospital Universitário Cassiano Antonio Moraes, Pronto Atendimento da Glória, Hospital Roberto Arnizaut Silvares) employees, nurses, doctors and patients that accepted to participate in this study. We would like to thank the Instituto Capixaba de Ensino, Pesquisa e Inovação em Saúde (ICEPi) and the Secretaria Estadual de Saúde do Espírito Santo (SESA) for clinical data of patients and RT-PCR results. This study was supported by FAPES (#151/2020) and CNPq (#401870/2020-0). We also thank Professor Edison Luiz Durigon and Dr. Danielle BL de Oliveira from Department of Microbiology at Institute of Biomedical Science (ICB) at University of São Paulo, USP – Brazil, for providing the inactivated SARs- CoV-2 virus strain (SARS.CoV2/SP02.2020.HIAE.Br GenBank accession number MT 126808.1).

## References

1. Li G, Fan Y, Lai Y, Han T, Li Z, Zhou P, et al. Coronavirus infections and immune responses. J Med Virol. 2020;92(4):424–432. doi: 10.1002/jmv.25685.

2. Khadke S, Ahmed N, Ahmed N, Ratts R, Raju S, Gallogly M, de Lima M, Sohail M. Harnessing the immune system to overcome cytokine storm and reduce viral load in COVID-19: a review of the phases of illness and therapeutic agents. Virology Journal 17(1) PMCID: PMC7558250.

3. Maffettone P, Oldani C. COVID-19: A Make or Break Moment for Global Policy Making. Global Policy DOI: 10.1111/1758-5899.12860 PMID: 32904957 PMCID: PMC7460972.

4. Bryant P, Elofsson A. Estimating the impact of mobility patterns on COVID-19 infection rates in 11 European countries. PeerJ 8:e9879.

5. Means AR, Wagner AD, Kern E, Newman LP, Weiner BJ. Implementation Science to Respond to the COVID-19 Pandemic. Front Public Health 8:462.

6. Vandenberg O, Martiny D, Rochas O, van Belkum A, Kozlakidis Z. Considerations for diagnostic COVID-19 tests. Nat Rev Microbiol.

7. Steel JJ, Sitko JC, Adkins MG, Hasstedt SC, Rohrer JW, Almand EA. Empowering academic labs and scientists to test for COVID-19. Biotechniques 69(4):245–248.

8. Pokhrel P, Hu C, Mao H. Detecting the Coronavirus (COVID-19). ACS Sens 5(8):2283–2296.

9. Zhou L, Chandrasekaran AR, Punnoose JA, Bonenfant G, Charles S, Levchenko O, Badu P, Cavaliere C, Pager CT, Halvorsen K Programmable low-cost DNA-based platform for viral RNA detection. Sci Adv 6(39).

10. Morel C, Lindahl O, Özenci V. Lessons from COVID-19 on the role of the state and the market in providing early testing. Journal of Global Health, 10(2).

11. Movasaghi Z, Rehman S, & ur Rehman DI (2008) Fourier Transform Infrared (FTIR) Spectroscopy of Biological Tissues. Applied Spectroscopy Reviews 43(2):134–179.

12. Baker MJ, et al. (2014) Using Fourier transform IR spectroscopy to analyze biological materials. Nat. Protocols 9(8):1771–1791.

13. Mitchell AL, Gajjar KB, Theophilou G, Martin FL, & Martin-Hirsch PL (2014) Vibrational spectroscopy of biofluids for disease screening or diagnosis: translation from the laboratory to a clinical setting. Journal of Biophotonics 7(3-4):153–165.

14. Freitas DLD, Câmara IM, Silva PP, Wanderley NRS, Alves MBC, Morais CLM, Martin FL, Lajus TBP, Lima KMG. Spectrochemical analysis of liquid biopsy harnessed to multivariate analysis towards breast cancer screening. Sci Rep 10(1):12818.

15. Martin FL, Kelly JG, Llabjani V, Martin-Hirsch PL, Patel II, Trevisan J, Fullwood NJ, Walsh MJ. Distinguishing cell types or populations based on the computational analysis of their infrared spectra. Nat Protoc 5(11):1748–1760.

16. Maitra I, Morais CLM, Lima KMG, Ashton KM, Date RS, Martin FL. Attenuated total reflection Fourier-transform infrared spectral discrimination in human bodily fluids of oesophageal transformation to adenocarcinoma. Analyst 144(24):7447-7456; PMID: 31696873.

17. Paraskevaidi M, Allsop D, Karim S, Martin FL, Crean S. Diagnostic Biomarkers for Alzheimer’s Disease Using Non-Invasive Specimens. Journal of Clinical Medicine DOI: 10.3390/jcm9061673 PMID: 32492907 PMCID: PMC7356561.

18. Araujo DB, Machado RRG, Amgarten DE, Malta FM, de Araujo GG, Monteiro CO, Candido ED, Soares CP, de Menezes FG, Pires ACC, Santana RAF, Viana AO, Dorlass E, Thomazelli L, Ferreira LCS, Botosso VF, Carvalho CRG, Oliveira DBL, Pinho JRR, Durigon EL. SARS-CoV-2 isolation from the first reported patients in Brazil and establishment of a coordinated task network. Mem Inst Oswaldo Cruz. 2020; 115:e200342; doi: 10.1590/0074-02760200342.

19. Corman VM, Landt O, Kaiser M, Molenkamp R, Meijer A, Chu DK, Bleicker T, Brünink S,Schneider J, Schmidt ML, Mulders DG, Haagmans BL, van der Veer B, van den Brink S,Wijsman L, Goderski G, Romette JL, Ellis J, Zambon M, Peiris M, Goossens H, Reusken C, Koopmans MP, Drosten C. Detection of 2019 novel coronavirus (2019-nCoV) by real-time RT-PCR. Euro Surveill 25(3); PMID: 31992387; PMCID: PMC6988269

20. Kennard RW & Stone LA (1969) Computer Aided Design of Experiments. Technometrics 11:137–148.

21. Lima KMG, et al. (2014) Classification of cervical cytology for human papilloma virus (HPV) infection using biospectroscopy and variable selection techniques. Analytical Methods 6(24):9643–9652.

22. Bro R, Smilde AK. Principal component analysis. Anal Methods 2014 6: 2812–2831.

23. Paraskevaidi M, Morais CLM, Lima KMG, Snowden JS, Saxon JA, Richardson AMT, Jones M, Mann DMA, Allsop D, Martin-Hirsch PL, Martin FL. Differential diagnosis of Alzheimer’s disease using spectrochemical analysis of blood. Proc Natl Acad Sci USA 114(38):E7929–E7938.

24. Lima KM, Gajjar KB, Martin-Hirsch PL, Martin FL. Segregation of ovarian cancer stage exploiting spectral biomarkers derived from blood plasma or serum analysis: ATR-FTIR spectroscopy coupled with variable selection methods. Biotechnol Prog 31(3):832-839; PMID: 25832726.

